# Acceptability of Rehabilitation Exoskeleton from the Perspective of Users with Spinal Cord Injury and Healthcare Professionals: a Mixed Methods Systematic Review

**DOI:** 10.1101/2024.09.18.24313846

**Authors:** Noémie Fortin-Bédard, Julien Déry, Margaux Simon, Andreanne K. Blanchette, Laurent Bouyer, Martine Gagnon, François Routhier, Marie-Eve Lamontagne

## Abstract

**Objective:** The objective was to document the acceptability of rehabilitation exoskeletons from the perspective of users with spinal cord injury (SCI) and healthcare professionals (HP).

**Methods:** This mixed-methods systematic review considered quantitative, qualitative and mixed methods studies that included adults with SCI using an exoskeleton for gait rehabilitation, as well as HP working within rehabilitation settings with individuals with SCI who used an exoskeleton. A convergent integrated approach per the Joanna Briggs Institute (JBI) was used.

**Results:** A total of 22 studies were included. Overall, individuals with SCI and HP expressed a favorable level of acceptability. Participants reported a positive affective attitude, an overall satisfaction, and several psychological benefits. Few burdens, ethical issues and opportunity costs have also been reported in the studies. Maintaining realistic expectations towards exoskeleton use and ensuring the appropriate selection of users is important for intervention coherence. In general, there was a positive perception regarding effectiveness and self-efficacy. Nevertheless, only a limited number of studies focused primarily on measuring acceptability, revealing an important gap in the literature.

**Conclusions:** The acceptability of exoskeletons among people with SCI and HP tends to be positive, which is promising for the sustainable implementation of this technology. However, there is still a lack of knowledge about the acceptability of HP, with only two studies conducted among this population. It is crucial to persevere in documenting the acceptability of exoskeletons, notably by standardizing comprehensive approaches for measuring acceptability, and to continue refining this technology.

## Introduction

In 2019, there were 9 million of people living with a spinal cord injury (SCI) around the world [1]. Living with an SCI affects physiological and psychosocial abilities, requiring considerable adaptation in daily life to these changes [2]. Resulting impairments and limitations, such as mobility, can reduce social participation and quality of life of these individuals [3]. Despite the rehabilitation process, most individuals with SCI live with persistent mobility limitations [4] That might vary according to the level and degree of the lesion, influencing the potential for walking.

In recent years, exoskeleton technologies have received significant media coverage as a novel and promising technology to support various human functions. An exoskeleton is an overground robotic device is an external structure placed along the segments that increases, aids or improves the user’s movement, locomotion, posture or physical activity [5]. While exoskeletons have primarily been utilized in the army and in research settings, they are implemented in healthcare settings around the world for the rehabilitation of people with SCI [6]. Emerging evidence supports that exoskeletons can also be used as movement-retraining devices to accelerate and increase the intensity of gait rehabilitation in people with SCI [7]. The use of exoskeletons thus presents potential benefits in the rehabilitation process, such as increased walking speed, improved balance and muscle strength, and reduced pain intensity and spasticity in people with SCI that persist even after the exoskeleton training is completed [8–12]. However, a substantial proportion of prior studies about the impact of exoskeleton use have adopted an exploratory approach. The additional value of exoskeleton use has not yet been consistently reported.

The rapid implementation of exoskeletons in healthcare settings highlights an urgent need to assess their acceptability. Acceptability is a multidimensional construct that has not been appraised consistently by researchers. Indeed, several definitions and theoretical or conceptual frameworks have been proposed to measure the determinants of acceptability of a technology, such as the Technology Acceptance Model (TAM) by Davis [13] and the Unified Theory of Acceptance and Use of Technology (UTAUT) by Venkatesh & Bala [14]. More recently, Sekhon et al. conducted an overview of systematic reviews of studies that aimed to define, theorize and measure the acceptability of healthcare interventions [15]. Then, a panel of experts reached a consensus defining acceptability as “a multi-faceted construct that reflects the extent to which people delivering or receiving a healthcare intervention consider it to be appropriate, based on anticipated or experienced cognitive and emotional responses to the intervention” [15]. This research team expanded the concept of acceptability by developing the Theoretical Framework of Acceptability (TFA) tailored to healthcare interventions [15]. This framework ensures the evaluation of intervention acceptability across three distinct timeframes such as pre-intervention (prospectively), during intervention (concurrently), and post-intervention (retrospectively), and considers the perspectives of both intervention providers and users [15]. The TFA facilitates the identification of key dimensions of an innovation that can be optimized to enhance its acceptability [16].

Despite growing interest in exoskeletons, several factors seem to influence their acceptability by users. There are two categories with distinct roles in the utilization, namely individuals with SCI, who wear the device, and healthcare professionals (HP), who operate it. On the one hand, various companies market several models of exoskeletons each with distinct intended purposes and characteristics [17]. These characteristics may possibly influence the perceived usability and ease of use, and consequently, the acceptability of the device [14]. On the other hand, the use and the acceptability of exoskeletons depend on the organizational context in healthcare settings (e.g., culture and values) as well as objectives and expectations regarding the use of this technology [18]. In this regard, a previous study identified that unmet expectations regarding the benefits associated with exoskeleton use was a barrier to device utilization [19]. The perceived acceptability is a key factor influencing the processes of implementation of a health intervention [15]. Indeed, previous studies highlighted that one of the major barriers to the use of a new technology, such as an exoskeleton, is low level of user acceptability [13]. Given that exoskeletons are a new technology increasingly implemented in health care facilities, such as rehabilitation facilities for people with SCI, it becomes imperative to focus on its acceptability to ensure the successful implementation.

Few studies have systematically appraised the acceptability when investigating the user experience of the exoskeleton. Nevertheless, some elements in these previous studies have been reported as determinants of the acceptability according to the TFA, such as ease of use and perceived benefits [20,21]. A comprehensive and integrated understanding of exoskeleton acceptability is currently lacking. There is a pressing need to synthesize existing knowledge and analyze individuals’ experiences using an acceptability framework. Since both quantitative (e.g., questionnaire of UTAUT by Venkatesh & Bala [14]) and qualitative (e.g., an interview) studies can shed light on the acceptability of an exoskeleton, a mixed method systematic review offers a comprehensive avenue to delve deeply into the experience of individuals with a SCI and healthcare professionals.

Considering the importance of user acceptability to promote the successful implementation of this technology, it is essential to conduct a knowledge synthesis. The objective of this systematic review was to document the acceptability of rehabilitation exoskeletons for individuals with SCI and healthcare professionals.

## Methods

The proposed mixed methods systematic review has been conducted in accordance with the Joanna Briggs Institute (JBI) methodology for Mixed Methods Systematic Review (MMSR) [22] and followed the PRISMA guideline [23]. A convergent integrated approach as defined by JBI (i.e., a combination of qualitative and quantitative data) was used to address the review question [22]. Protocol was registered in PROSPERO (CRD42023401829).

### Search Strategy

A three-step search strategy was used in this review supervised by a librarian specialized in systematic reviews (M.G.). An initial limited search of Medline (via Ovid), and CINAHL (via EBSCO) databases was undertaken followed by an analysis of the text words contained in the title and abstract and the index terms used to describe the articles to identify the keywords. The search strategy, including all identified keywords and index terms was adapted for each included information source on March 23, 2023. The databases that were searched included Medline (via Ovid), CINAHL (via EBSCO), Embase, and Web of Science databases. These databases were chosen since they are the main sources of articles in the fields of medicine and rehabilitation, with Web of Sciences being selected as a multidisciplinary database. The full search strategy is provided in Supplemental material Table 1.

**Table 1.**
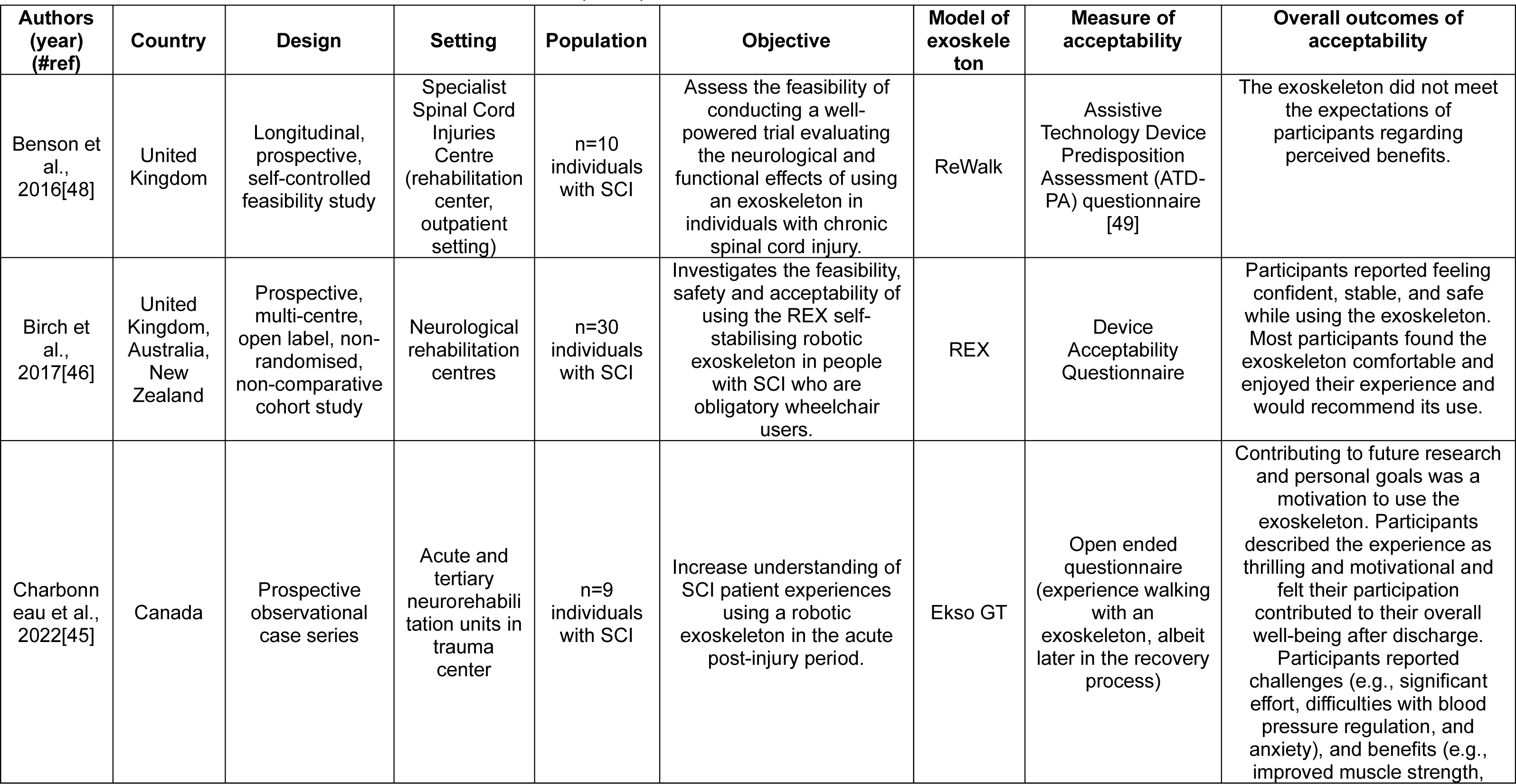
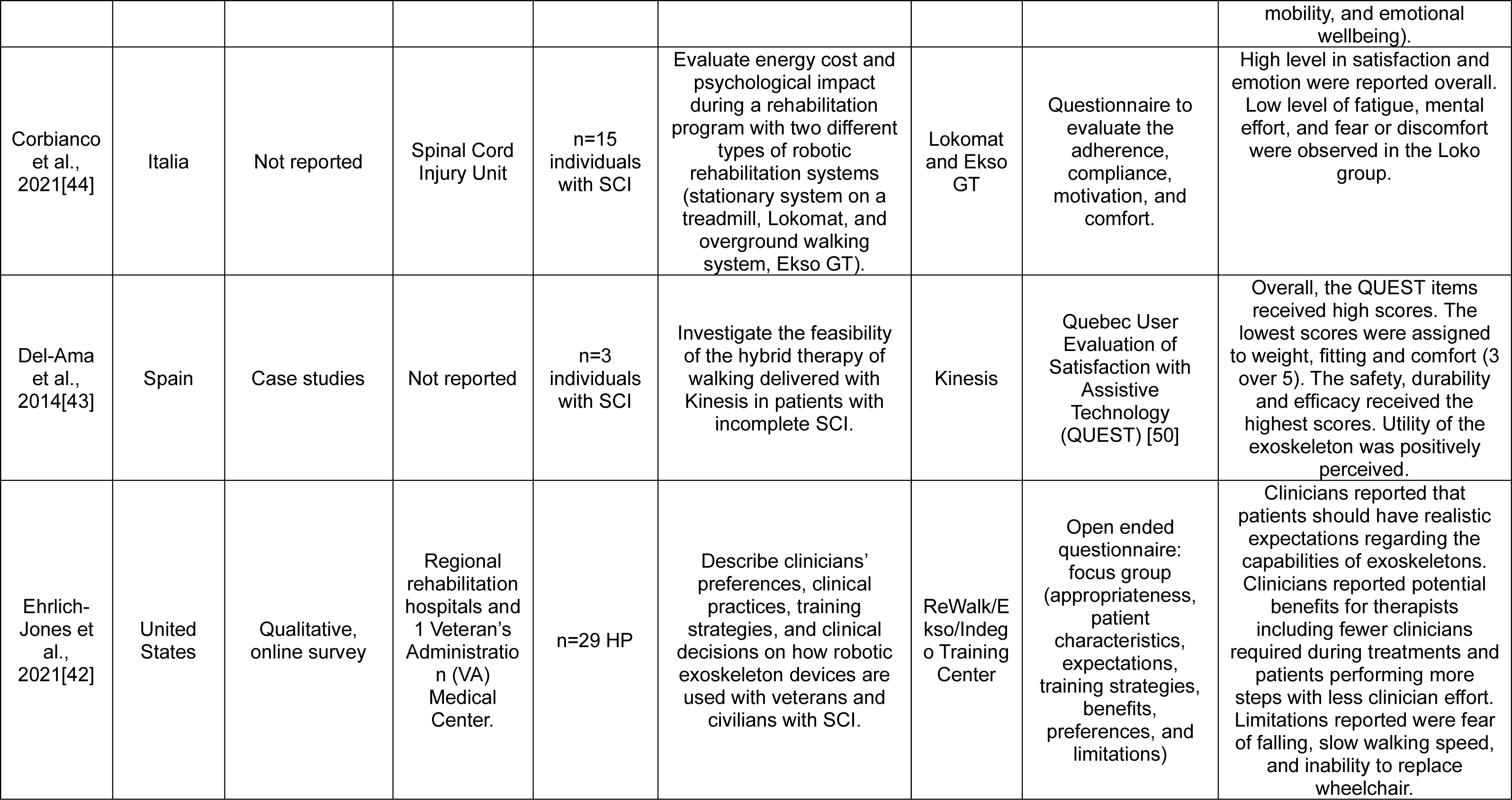
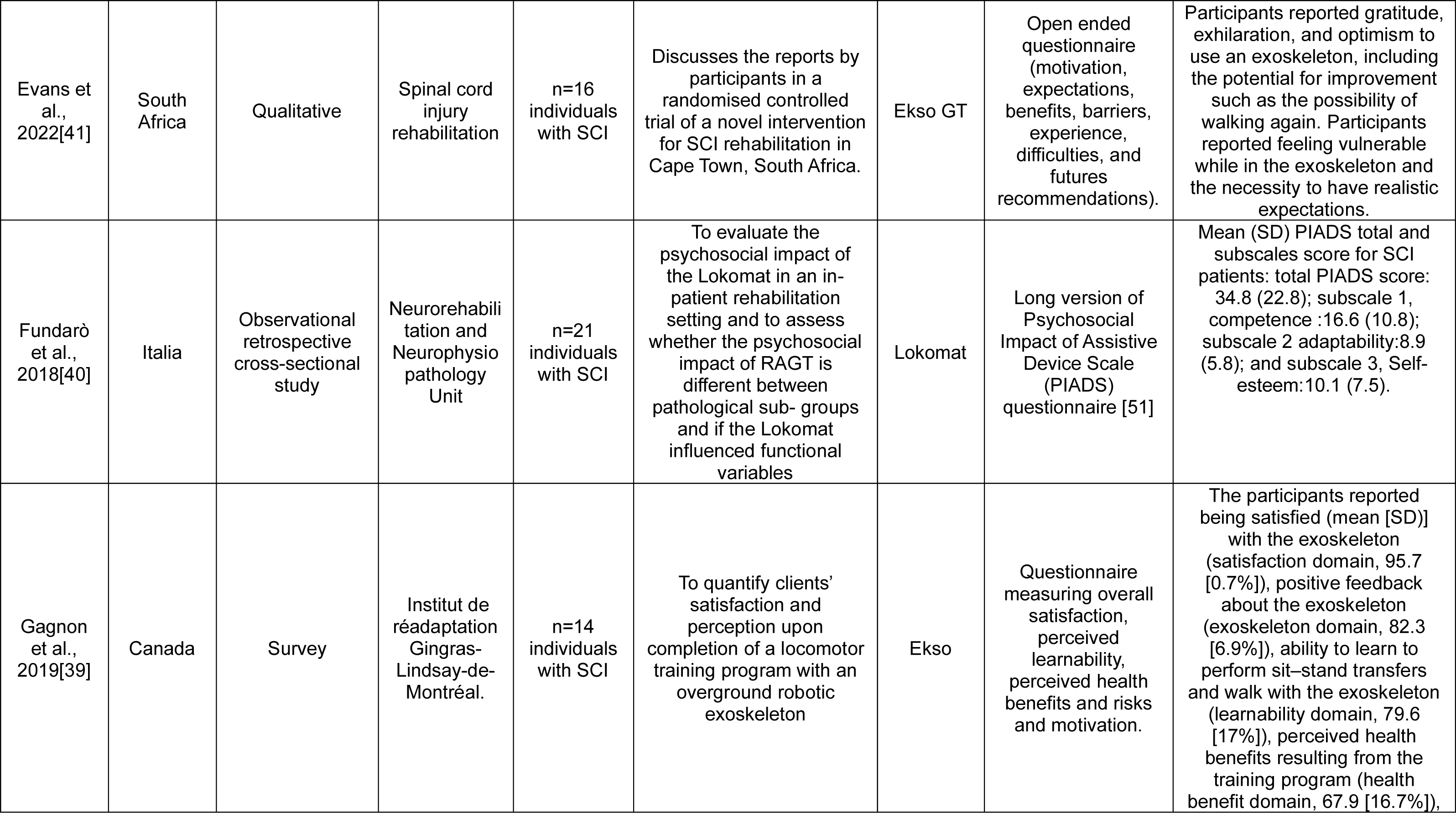
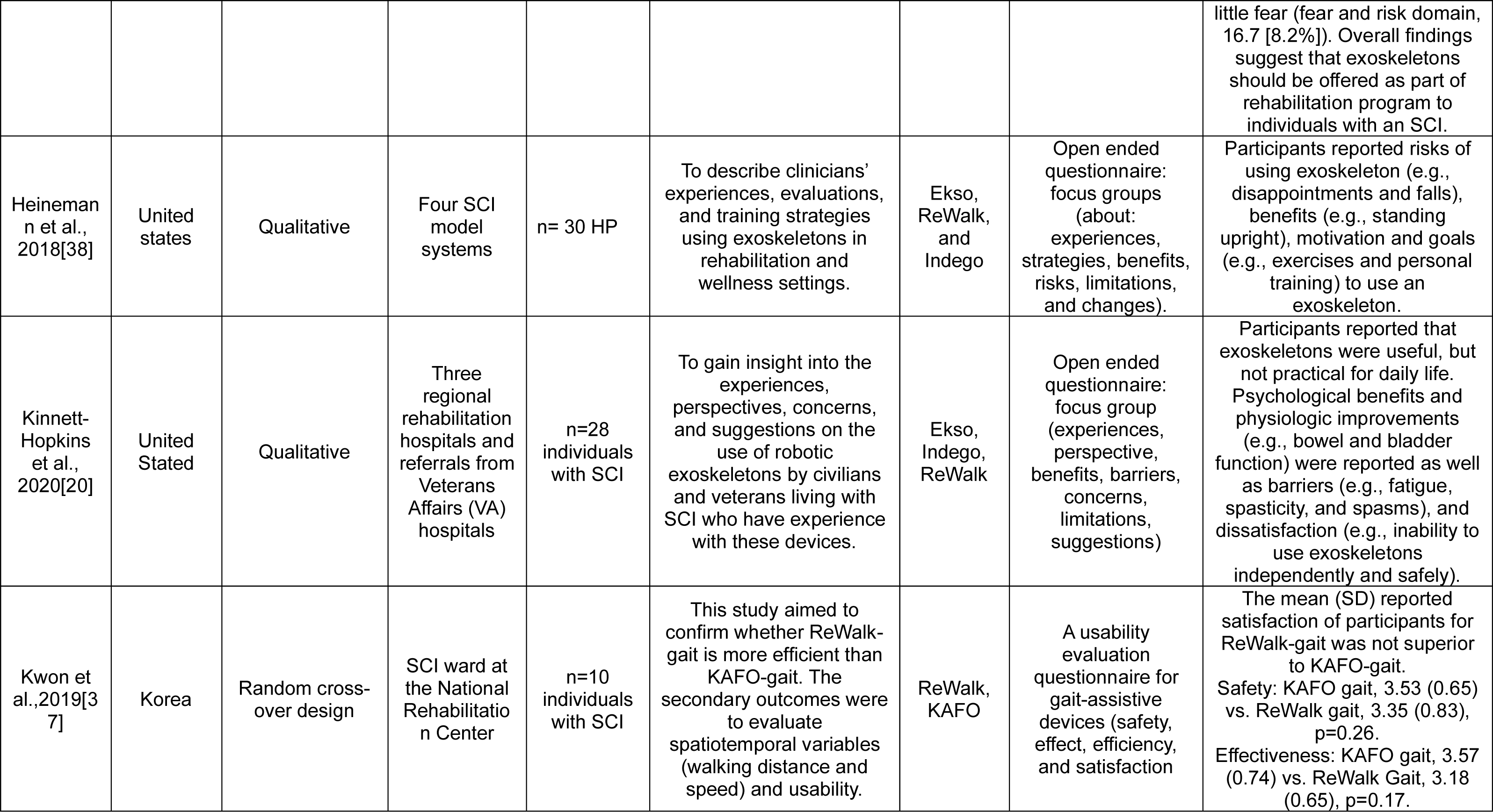
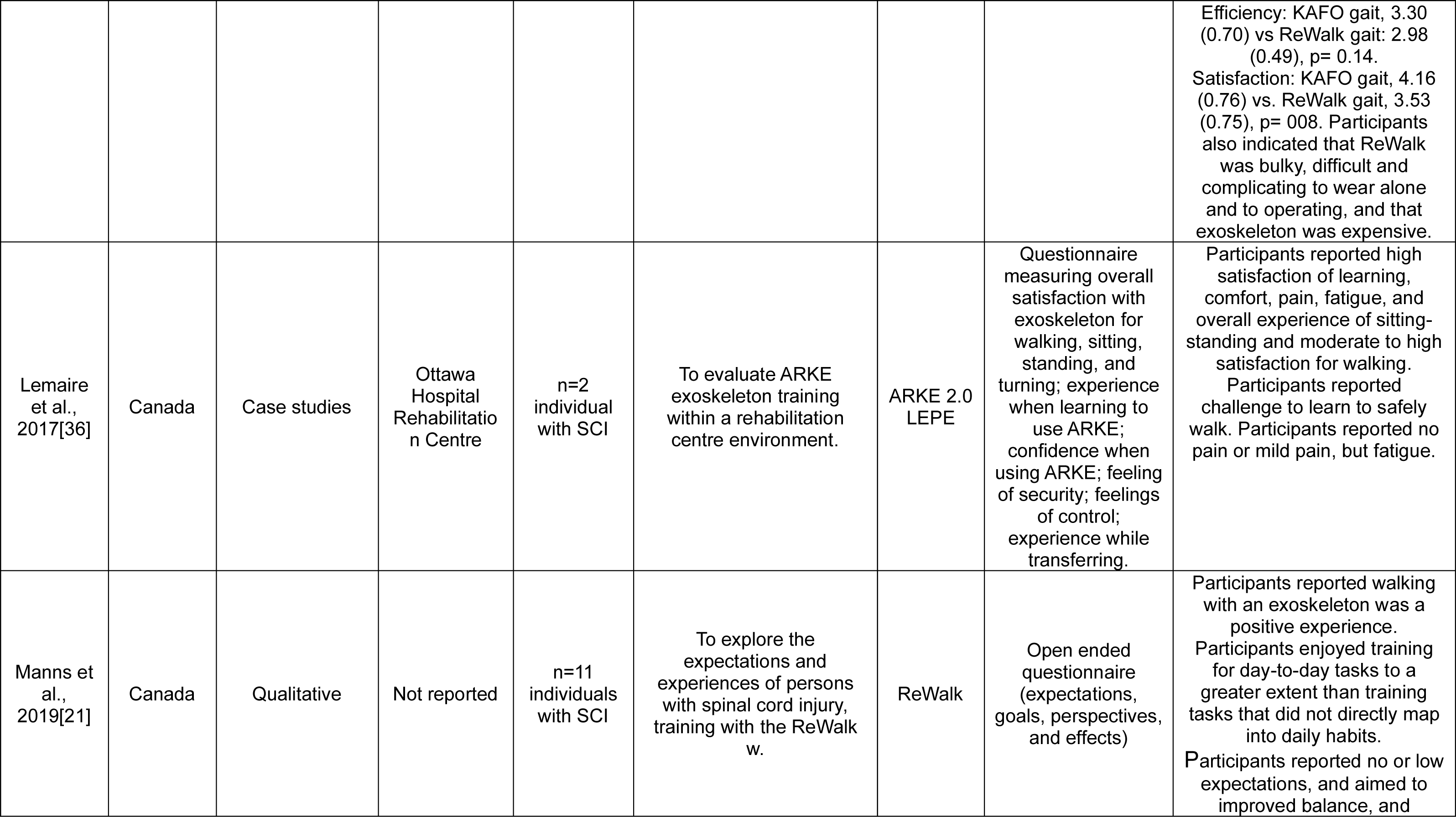
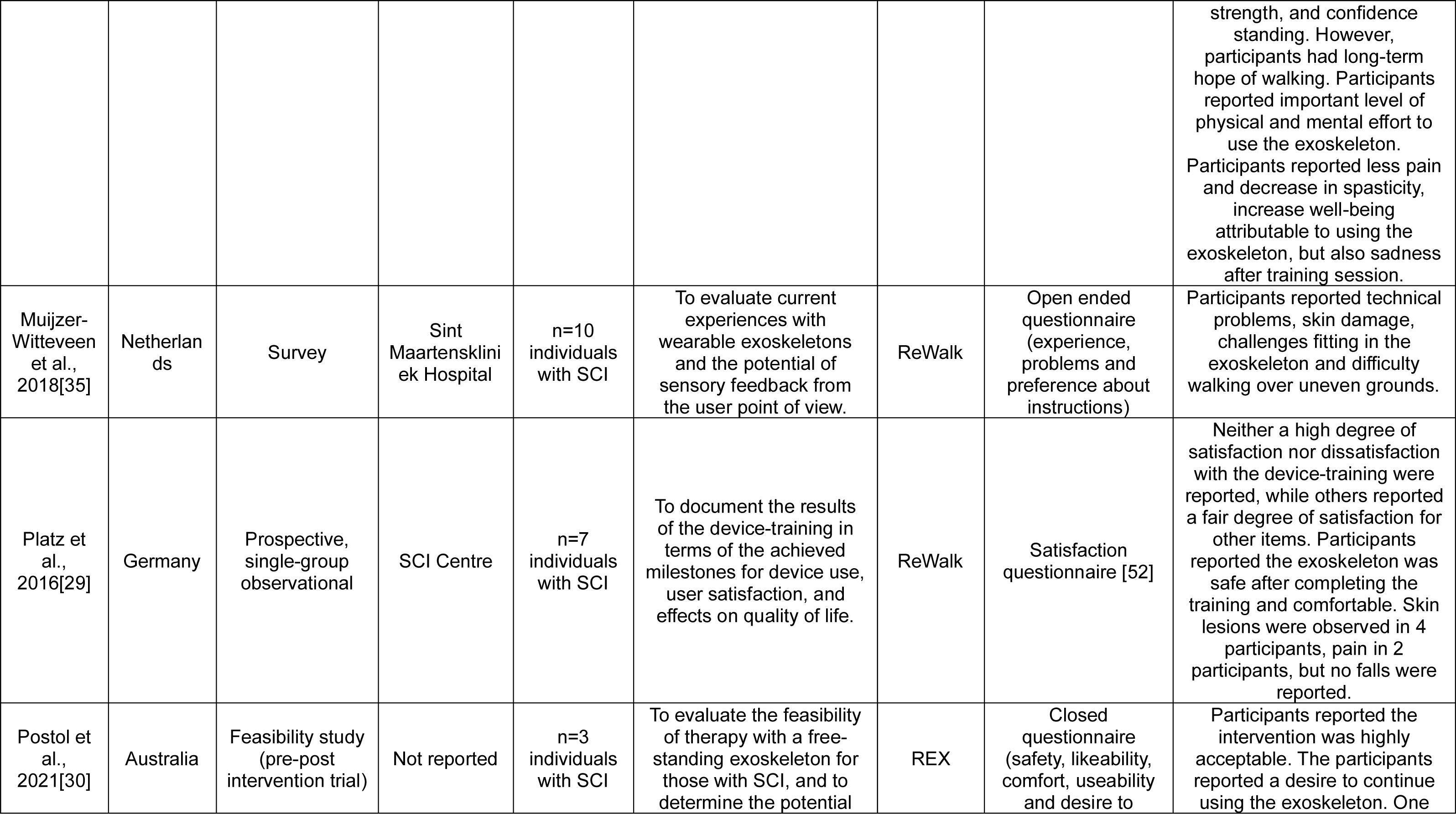
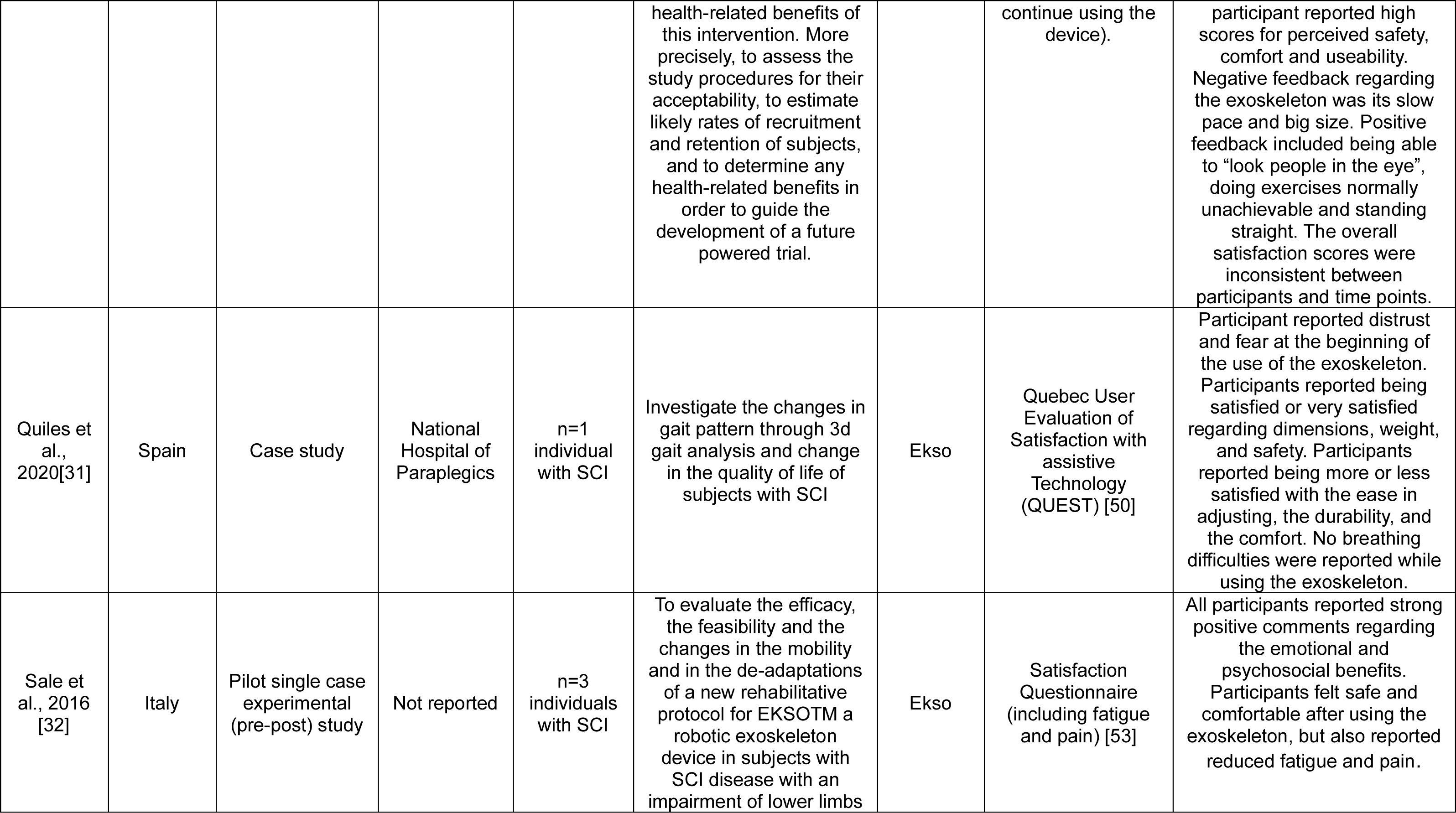
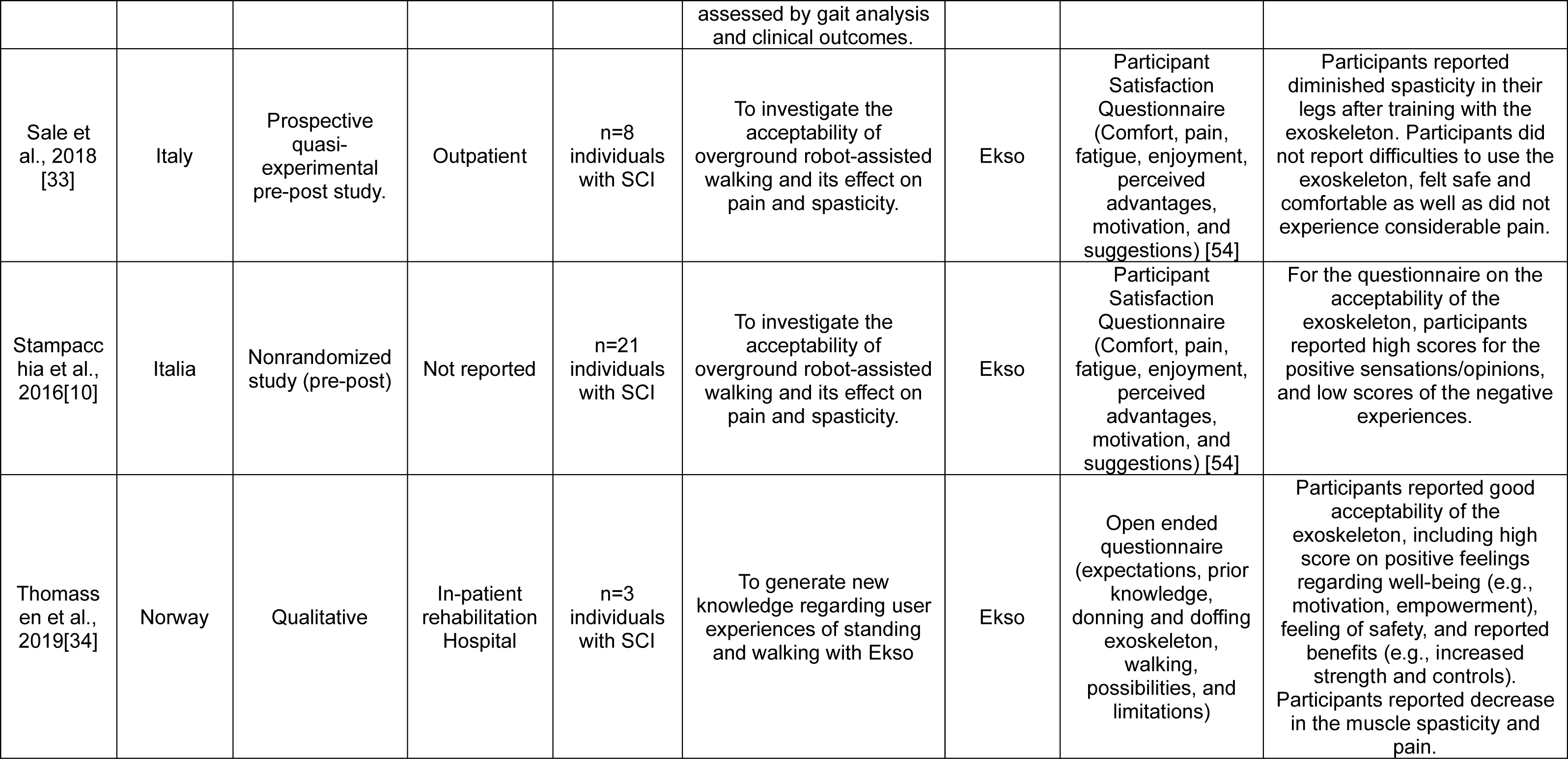
Characteristics of included studies (n=22).

### Eligibility Criteria

This review considered quantitative, qualitative and mixed methods studies that included adults aged 18 years and older with SCI who used an exoskeleton for gait rehabilitation, and studies involving HP in rehabilitation settings using exoskeleton with SCI patients. Searches were limited to publications in English and French, as members of the research team speak both languages, without any restrictions related to the publication date.

Studies documenting the acceptability of rehabilitation exoskeletons in term of perspective among individuals with SCI and HP who used an exoskeleton as part of rehabilitation were included. The reported results must relate to at least one construct of the theoretical framework of acceptability (TFA) among the seven component constructs (i.e., affective attitude, burden, perceived effectiveness, ethicality, intervention coherence, opportunity costs, and self-efficacy) [15].

This review excluded: 1) studies that focus on personal, at-home or in-the-community use of an exoskeleton, 2) the following types of articles or study design: conference proceedings, commentaries, letters, book chapters, animal studies, editorials, reviews and meta-analyses, abstracts, proof-of-concept; and 3) studies focusing on outcomes related to function, design and clinical effectiveness of an exoskeleton without documenting user acceptability to understand the perspective of users. These studies were excluded because exoskeletons used as technical aids in a personal setting or at home do not have the same characteristics, nor the same objectives of use than exoskeleton used in gait rehabilitation. Both of which could influence user acceptability.

### Screening and Selection Process

After conducting the literature search, all identified studies were gathered and uploaded into Endnote 20 (Clarivate Analytics, PA, USA) with duplicates subsequently removed [24]. Titles and abstracts were then screened by two independent reviewers (N.F.-B. and J.D.) to assess if the inclusion criteria were met for the review using Covidence software platform [25]. The full texts of selected studies were retrieved and assessed in detail against the inclusion criteria by two independent reviewers (N.F.-B. and J.D.). Full-text studies that did not meet the inclusion criteria were excluded and reasons for exclusion are provided in Figure 1. Any disagreements that arose between the reviewers were resolved through discussion with a third reviewer (M-E.L.). Cohen’s kappa coefficient during the title and abstract screening was 0.66, and 0.52 during the full text screening, suggesting substantial and moderate agreement, respectively [26].

**Figure 1.**
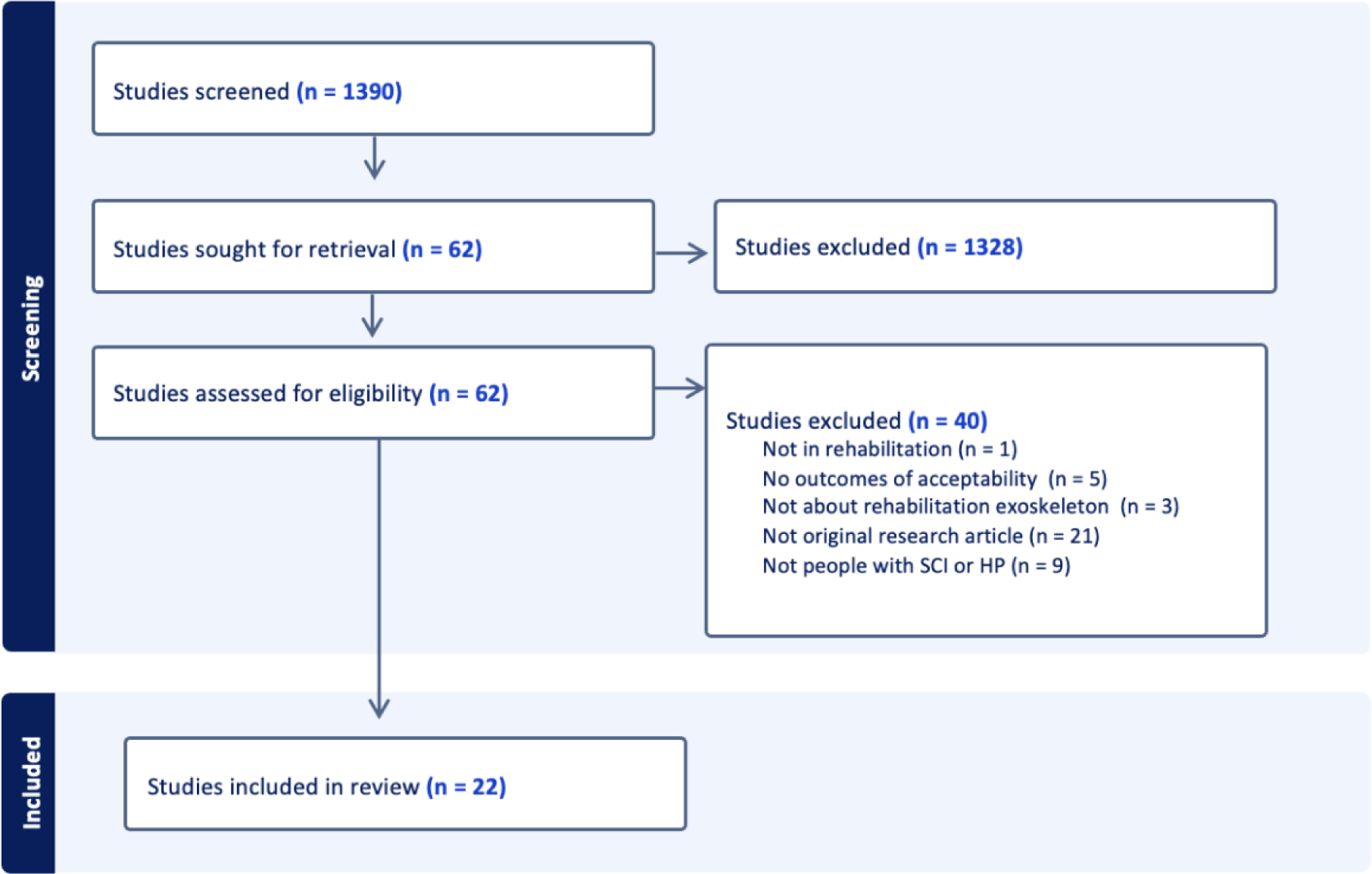
PRISMA flowchart

### Data Extraction

Quantitative and qualitative data were extracted from included studies by two independent reviewers (N.F.-B. and J.D.) using Microsoft Excel (See Table 1). The data extracted included title, author, years of publication, country, study design, setting, objective of the study, details about the population, eligibility criteria, dropouts and related reasons, model of exoskeleton, context of use, main measures, measure of acceptability, overall acceptability, and time points acceptability. Any discrepancies between the reviewers in the extracted data were resolved through discussion, or with the involvement of a third reviewer (M-E.L).

### Data Synthesis and Integration

The convergent integrated approach according to the JBI methodology for mixed methods systematic review was used in this review [22]. This involved assembling the “qualitized” data with the qualitative data [22]. This involved the transformation of the quantitative results into textual descriptions or narrative interpretation to allow integration with qualitative data [22]. Assembled data were categorized and pooled together, in the software Nvivo 14 [27], using a deductive approach from the Theoretical framework of acceptability (TFA) [15]. Data were categorized according to the seven component constructs of TFA, namely 1) affective attitude; 2) burden; 3) perceived effectiveness; 4) ethicality; 5) intervention coherence; 6) opportunity costs, and 7) self-efficacy [15].

### Critical Appraisal

Eligible studies underwent critical appraisal by two independent reviewers (N.F.-B., and J.D.) for methodological quality using the Mixed Methods Appraisal Tool (MMAT) [28]. This tool enables the assessment of the quality of qualitative, quantitative and mixed methods studies in conducting a sensibility analysis according to evaluation criteria rather than calculate an overall score [28]. Any disagreements between the reviewers were resolved through discussion with a third reviewer (M-E.L).

## Results

A total of 2338 studies were extracted from the four databases, and among these, 948 were removed manually or by Covidence due to duplicates. Thus, 1390 studies were screened for titles and abstracts and 62 studies were subsequently assessed in full text. Of these, 40 were excluded because they were not conducted in a rehabilitation setting, did not focus on acceptability outcomes, did not involve the use of a rehabilitation exoskeleton, were not original research articles, or were not among people with SCI or HP. Finally, 22 studies were included in our review [10,20,21,29–47].

### Characteristics of Included Studies

Detailed characteristics of the included studies can be found in Table 1.

The studies have been published between 2014 and 2022. Only three studies (14%) explicitly had as main goal to measure the acceptability of an exoskeleton [30,31,46], while the remaining studies solely reported certain domains of acceptability. The majority of studies documented the perspective of individuals with SCI (i.e., those receiving the intervention) [10,20,21,29–37,39–41,43–46,48], whereas only two studies (9%) explored the acceptability of the exoskeleton from the HP standpoint (i.e.. those delivering the intervention) [38,42], representing a total of 216 individuals with SCI and 59 HP. The Ekso exoskeleton (including Ekso GT) was the most commonly utilized in the included studies [10,32–34,39,41,45], followed by the ReWalk, which was the second most used exoskeleton [21,29,35,48]. Some studies, conducted in rehabilitation centers, also included more than one model of exoskeleton (i.e., ReWalk, Ekso, Indego, Lokomat and/or KAFO) [20,37,38,42,44]. Other models of exoskeletons, such as REX [46], Kinesis [43], Lokomat [40], ARKE 2.0 LEPE [36], and H2 portable [31] lower limb exoskeleton were used in the studies included.

The timing of the acceptability assessment regarding the use of the exoskeleton varies among studies. Most studies (n=15; 68%) measured acceptability at only one time point, either during [44] or after the use of the exoskeleton [10,20,29,31,34–40,42,43,45,46]. Two studies measured acceptability before, prospectively and retrospectively the use [21,41], while two other studies (9%) assessed acceptability concurrently and retrospectively [32,33]. Finally, only two (9%) studies conducted assessments of acceptability at three timepoints, i.e., prospectively, concurrently, and retrospectively [30,48].

Five studies (23%) were conducted in Italy [10,32,33,40,44], four (18%) in Canada [21,36,39,45], three (14%) in the United States [20,38,42], and two studies in Spain (9%) [31,43]. Only one study (5%) is multicentric, including the United Kingdom, Australia, and New Zealand [46]. Finally, studies have been conducted in other countries including the United Kingdom [47], South Africa [41], Korea [37], the Netherlands [35], Germany [29], Australia [30], and Norway [34].

### Methodological quality

The evaluation of the quality of quantitative, qualitative, and mixed methods studies included in this review are available in Supplemental (Table 2). There were seven qualitative studies (32%) [20,21,34,38,41,42,45] included in the present review. Overall, qualitative studies were of good quality, with an adequate sample size and sufficiently detailed data. For example, Charbonneau et al., 2022, presented a reflexivity statement [42,45]. Ten studies (45%) were non-randomized quantitative studies [10,29,30,33,37,40,43,44,46,48]. The latter encompassed small samples ranging from one participant to thirty participants (e.g., [43,48]), with the majority failing to consider confounding variables in the study design or analysis of results (e.g., severity and level of SCI or time since injury). Finally, six studies (27%) were descriptive quantitative studies [31–33,35,36,39]. As in the non-randomized studies, some of the measurement tools used to assess domains of acceptability were not gold standards, or an appropriate rationale for the choice of measure was not provided. In addition, some studies omitted the presentation of inclusion criteria, as well as the details concerning dropouts and their associated reasons.

### Findings of the review

Results were integrated into the seven domains in coherence with the TFA definitions [15]. Main results are presented according to these seven domains in Figure 1 (which summarizes the main results and definition of domains [55]) and detailed accordingly in the following subsections.

**Figure 1.**
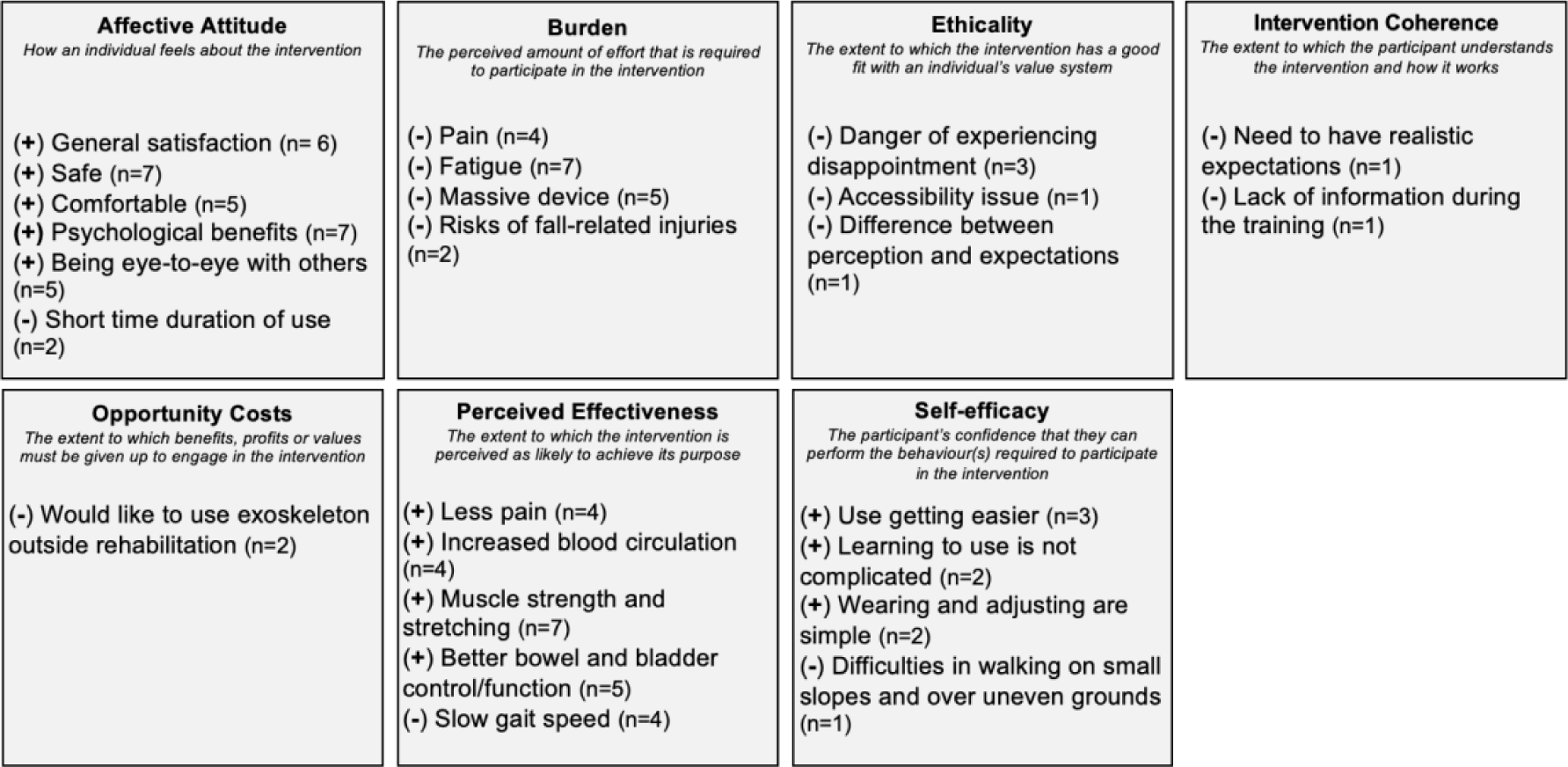
Summary of the main results.

### Affective attitude

Six studies (27%) stated that people with SCI were generally satisfied with the use of the exoskeleton [10,30,32,39,40,44]. Among these studies, only three (14%) [10,30,40] had as main their objective to measure the acceptability of this technology, describing it as acceptable. However, satisfaction of the use of the exoskeleton varied in other studies [20,30]. For example, Kinnett-Hopkins et al., 2020 [20] highlighted that a few participants reported dissatisfaction with the technology, particularly because they could not use it independently at home and feel safe. Expectations and goals for the use of the exoskeleton, when reported, were different among the studies. Indeed, two studies (9%) reported that the reason to participate in a research study and using an exoskeleton was to benefit other people in their rehabilitation in the future [21,45]. Interestingly, two studies (9%) reported that participants participated in research projects involving the use of an exoskeleton with the goal of walking during rehabilitation [20,45]. Three studies (14%) reported that not all initial expectations regarding the use of an exoskeleton in rehabilitation were met [35,42,48].

Participants of seven studies (32%) reported feeling safe while using the exoskeleton [29,32–34,36,43,46]. In contrast, four studies (18%) reported a fear of falling [21,31,41,42] and one of them further specified that this fear of falling occurred especially at the beginning of the training with the exoskeleton [31]. In addition, in five studies (23%), participants reported that the exoskeleton was comfortable [29,32,33,39,46]. However, participants in the study of Quiles et al., 2020 reported being neutral with the comfort of the exoskeleton [31]. Thomassen et al., 2019 specified that the feeling of safety was due to being well strapped to the exoskeleton [34].

Seven studies (32%) [20,32,34,36,39,41,45] have reported various psychological benefits and positive impacts on the well-being of people with SCI who have used an exoskeleton during their rehabilitation. Empowerment, motivation, and increased self-confidence were positive feelings reported [34,45]. In addition, being at the same eye level as others [20,21,30,39,45], the positive experience of standing and gives the sensation of walking again [20,30,34,38] and feeling liberated by the open space around them [34]. However, two studies (9%) [34,45] have highlighted feelings of frustration and disappointment notably due to the short-lasting positive sensations related to the use of the exoskeleton (e.g., better control of movements and gastrointestinal function). Participants would have liked to use the exoskeleton for a longer period to optimize the effects [21]. Finally, the feeling of being completely locked into the device and the unnatural feeling when wearing the device was reported in one study [34].

### Burden

Among the studies reporting dropouts and their reasons, skin breakdown problems, talus fracture, not enjoying the device, fear, or time restraints [31,43,48] were reported. Seven studies (32%) noted that people with SCI felt tired during or after the use of an exoskeleton due to high physical and cognitive demands (e.g., concentration, cognitive exertion, and mental effort) [20,21,29,32,34,36,39,45]. Two studies (9%), including one conducted among HP, reported the risks of fall-related injuries for individuals with SCI [20,38]. In addition, five studies (22%) reported that the exoskeleton was too massive [20,21,30,34,37]. While participants in three studies (14%) [29,32,33] reported that the use of the exoskeleton did not result in significant pain, four other studies (18%) reported pain [29,34,36,45], potentially leading to unpleasant training experiences [45]. Finally, the high cost of the exoskeleton [20,37], technical bugs [35] of equipment to use the exoskeleton and skin damage have been reported [29,35]. HP also reported the demand for support and training necessary to use the device [38,42]

### Ethicality

Few studies have reported on ethical issues surrounding the use of exoskeletons. However, three studies (18%) reported the danger of experiencing considerable disappointment following false hope for people with SCI using an exoskeleton, such as walking independently [20,38,41]. Evans et al., 2022 reported that this could be particularly present in the context of poor access to services [41], in addition to accessibility issues for an exoskeleton [38]. Moreover, Kinnett-Hopkins et al., 2020 reported potential ethical issues due to the difference between the perception and expectations of able-bodied individuals toward exoskeletons that could differ from those of people with SCI [20]. More precisely, people with SCI may be pressured to conform to able-bodied normative views to an extent grater to the actual benefits of the exoskeleton, which could lead to unrealistic expectations.

### Intervention Coherence

According to clinicians in the study of Ehrlich-Jones et al., 2021, people with SCI using an exoskeleton in rehabilitation need to have realistic expectations toward the outcomes of the device use to maintain intervention coherence and to avoid being disappointed [42]. In this regard, the appropriateness of the use of the exoskeleton to ensure intervention coherence depends, among other things, on patient goals [42]. Moreover, participants with SCI in the study of Muijzer-Witteveen et al., 2018 reported a lack of information regarding how to use the exoskeleton during the training. [35].

### Opportunity Costs

No study has clearly indicated that benefits, profits, or values, such as other valuable rehabilitation activities, have to be given up in order to use an exoskeleton. Nevertheless, even if participants of the study by Manns et al., 2019 enjoyed their experience while using this technology, they noted, as a limitation, that the exoskeleton could not be used outside of rehabilitation settings. In this regard, Kinnett-Hopkins et al., 2020 reported that the exoskeleton had limited use outside rehabilitation [20]. Thus, the use of exoskeletons is limited to a rehabilitation context and the ability to use it cannot be transferred to real-life context.

### Perceived Effectiveness

Studies reported various perceived improvements in impairments and physical benefits of the use of an exoskeleton in individuals with SCI [39,45], such as increased muscle strength [20,21,34,39,41,45], improved bowel and bladder function [20,21,38,41,45], increased blood circulation [21,34,41,45] and feeling warmth [21,34]. In addition, participants of four studies (18%) have reported experiencing reduced pain or improved pain management while using the exoskeleton [20,21,38,45]. However, Manns et al., 2019 specified that pain came back two months after the end of the training with the exoskeleton [21]. Studies reported mixed results regarding self-reported improvement in spasticity among participants. Indeed, four studies indicated improvement while one study reported no change [20,21,32,33,41]. HP highlighted various factors they perceived as influencing the successful use of the device including motivation, general health, learning style, confidence, and body awareness [42]. Despite the exoskeleton’s effectiveness, participants in four studies have noted specific limitations of the device, particularly a slow walking speed [20,30,34,42].

### Self-Efficacy

Three studies (18%) reported that the use of an exoskeleton improved over time (i.e., more difficult at the beginning but less at the end) [20,21,45]. Participants from two studies (9%) reported feeling confident while using an exoskeleton [29,46]. Participants from Sale et al., 2016 and Sale et al., 2018 indicated that learning to use the device was not complicated [32,33]. Furthermore, wearing and adjusting the device was simple [32,33]. However, other studies reported various experiences, such as difficulty to transfers in exoskeletons [34], and difficulty to wear and operating it alone [37]. Lemaire et al., 2017 also reported challenge to learn to safely walk with the exoskeleton [36]. In addition, participants in the study of Muijzer-Witteveen et al., 2018 reported difficulties with determining the body position in addition to difficulties in walking on small slopes and walking over uneven grounds [35]. Finally, participants in Gagnon et al., 2019 also reported that it was easy to perform sit-stand transfers [39].

## Discussion

The objective of this mixed methods systematic review was to document the acceptability of rehabilitation exoskeletons for individuals with SCI and HP. Overall, the acceptability of exoskeletons from the perspective of people with SCI and HP was generally positive. However, only two studies focused on acceptability of HP, limiting the generalization of this conclusion among this population. A positive affective attitude was reported with good general satisfaction and several psychological benefits. There was a generally positive trend in perceived effectiveness and self-efficacy. In addition, few burdens, ethical issues and opportunity costs have been reported in the present review. Therefore, future studies should focus on these factors to better understand the acceptability of this device. Plus, only three studies aimed to measure acceptability as their primary objective, which highlights an important gap in the literature [30,31,46].

The results of the present synthesis are overall consistent with previous studies documenting the acceptability of health intervention using technologies in individuals with SCI. For example, studies examining the use of teleconference for testing cognitive abilities, or hand-cycling high-intensity interval training for people with SCI identified positive acceptance [56,57], potentially indicating a good technology acceptance from this population. Thus, the present findings contribute to the understanding that individuals with SCI perceive technologies used in their rehabilitation as acceptable. However, as with the use of exoskeletons, it is important for the intervention to maintain realistic expectations of technology use and to ensure the selection of the right user. These considerations are also important for the use of other technologies in rehabilitation, such as virtual reality [58].

Individuals having regular contact with HP or other individuals with SCI can also influence the acceptability of the device and explain the positive acceptability as reported by Evans et al., 2022 and Manns et al., 2019 [21,41]. Indeed, a previous study highlighted the positive impact of professional and peer support during the rehabilitation process [59]. Also, determining whether exoskeletons are considered equally acceptable by HP as by people with SCI is difficult, since only two studies involving HP were included in this review. Considering the important role of HP in initiating the use of an exoskeleton in rehabilitation, more studies are needed to explore the acceptability of exoskeletons from the HP’s point of view. In addition, future studies should be carried out using a qualitative approach using a conceptual framework to explore acceptability [60]. Indeed, this will allow for an in-depth and systematic exploration of the complexity of the acceptability of this innovative and evolving technology. Finally, some exoskeletons have been studied more than others according to the included studies (e.g., Ekso and ReWalk), and future studies should be conducted on various exoskeletons, considering their specific characteristics and designs.

In the present review, factors limiting the acceptability of exoskeletons included, for example, the bulkiness of the device, pain, and technical problems. Similarly, others health technologies, such as artificial intelligence and functional electrical stimulation in physical rehabilitation have reported preoccupation regarding technological problems (e.g., battery lifetime) [61,62]. These issues decreased usability and potentially cause frustration among users, leading to interruptions in care, which could negatively influence the acceptability of the technology [62,63]. Considering the importance of the device and intrinsic characteristics per se in determining its acceptability, suggestions for manufacturers to improve their devices have been reported in previous studies. These potential improvements include characteristics such as a lighter weight, a system to reduce fall risk, adding the capacity to ascend and descend stairs with the exoskeleton and being more adjustable [20,34,38,42]. To facilitate adoption and implementation of exoskeletons in rehabilitation, users must be involved right from the start of the development and all implementation stages [64]. Furthermore, studies in the present review included relatively homogeneous samples of individuals with complete and incomplete SCI considering the eligibility criteria for the use of an exoskeleton. Consequently, elderly people, people with a larger body mass index and children were never eligible in the included studies. In this regard, Postol et al. reported that one of the main reasons for the exclusion of potential participants was due to their physical characteristics (i.e., weight and height) that did not fit with the device [30].In addition, participants in the studies were not distinguished based on their prognosis (e.g., potential for walking). There may be variations in expectations regarding exoskeletons, which would be interesting to investigate in the future. To ensure increased acceptability and equity for all, greater adaptability of the technology to various individuals with SCI and context will be crucial, in addition to an unequivocal demonstration of the added value of using an exoskeleton in rehabilitation. Besides, according to the Consolidated Framework for Implementation Research (CFIR), adaptability is an important criterion for successful implementation [65].

Numerous methodological factors may have influenced the results of studies included in the study, and consequently, the present knowledge synthesis. First, social desirability biases, especially in the case of an innovative technology with limited access [66], may influence the acceptability of participants. Moreover, there may be selection biases. For example, many of the studies included in this review did not report dropout rates and reasons for dropping out, which would have provided a better understanding of acceptability. Third, various tools and approaches were used to measure acceptability concepts among the included studies. For example, two studies used the Quebec User Evaluation of Satisfaction with Assistive Technology (QUEST) [31,43], while this tool only measures certain aspects of acceptability, i.e., satisfaction toward assistive technology [67]. More precisely, opportunity costs and ethical issues are important aspects of acceptability which have not been widely discussed in the literature to date making it difficult to provide overall acceptability results [15]. Considering that the exoskeleton is a rapidly evolving technology that is increasingly implemented in rehabilitation settings, future studies should use a uniform and comprehensive measurement tool for acceptability to observe how its acceptance grows in various users.

To the best of our knowledge, this systematic review is the first to incorporate a theoretical framework of acceptability to combine the results of previous studies. Another strength of our synthesis is the comprehensive search strategy that included both quantitative, qualitative, and mixed methods studies to explore acceptability in depth. However, the limitations of our work must also be addressed. First, we experienced challenges in categorizing the study’s results across the seven domains of the TFA, given the broad definitions of these domains. However, the analysis was conducted by two independent reviewers, and a third person was available in case of disagreements, favoring the mutual exclusion of the domains. For example, one reviewer considered a result to be related to the “Opportunity cost” construct, while another reviewer considered it to be related to the “Perceived Effectiveness” construct. To resolve this conflict, the reviewers consulted the third independent reviewer (M-E. L) to decide while referring to the definition of each construct. Second, the included studies involved small samples of users and validated questionnaires were rarely utilized, consequently constraining the reproducibility of the studies. In turn, these limitations limit the generalizability of the conclusion of the present study. Third, the MMAT quality appraisal tool items are subject to interpretation and less objective [68]. Finally, we were unable to conduct additional searches on the Scopus multidisciplinary database, as our university does not subscribe to this database.

## Conclusions and Recommendations

The perspective of exoskeletons appears favorable among individuals with SCI and HP. This acceptability is promising for the sustainable implementation of this device. However, it is crucial to continue research efforts into the acceptability of exoskeletons, notably by standardizing ways of comprehensively measuring acceptability, and to further develop this technology for optimal use and benefit of people with SCI. There is also a need to better document the acceptability of HP.

## Data Availability

All data produced in the present work are contained in the manuscript

## Acknowledgments

N.F-B was supported through a scholarship from the Réseau provincial de recherche en adaptation-réadaptation (REPAR) and hold a master training award from the Canadian Institutes of Health Research (CIHR) and from the Quebec Health Research Funds (FRQS) (# 311138) in partnership with the Unité de soutien au système de santé apprenant (SSA) Québec. J.D was supported through a scholarship from FRQS. M.S was supported through a scholarship from the Centre interdisciplinaire de recherche en réadaptation et intégration sociale (Cirris). Salary supports for F.R were provided by the FRQS Senior Research Scholar program.

## References

1. Liu Y, Yang X, He Z, et al. Spinal cord injury: global burden from 1990 to 2019 and projections up to 2030 using Bayesian age-period-cohort analysis [Review]. Frontiers in Neurology. 2023 2023-December-05;14.

2. Barone SH, Waters K. Coping and Adaptation in Adults Living With Spinal Cord Injury. Journal of Neuroscience Nursing. 2012;44(5).

3. Noreau L, Noonan VK, Cobb J, et al. Spinal cord injury community survey: a national, comprehensive study to portray the lives of canadians with spinal cord injury. Top Spinal Cord Inj Rehabil. 2014 Fall;20(4):249–64.

4. Franceschini M, Di Clemente B, Rampello A, et al. Longitudinal outcome 6 years after spinal cord injury. Spinal Cord. 2003 2003/05/01;41(5):280-285.

5. Lowe BD, Billotte WG, Peterson DR. ASTM F48 Formation and Standards for Industrial Exoskeletons and Exosuits. IISE Transactions on Occupational Ergonomics and Human Factors. 2019 2019/10/02;7(3-4):230–236.

6. Forte G, Leemhuis E, Favieri F, et al. Exoskeletons for Mobility after Spinal Cord Injury: A Personalized Embodied Approach. Journal of personalized medicine. 2022;12(3).

7. Grasmücke D, Zieriacks A, Jansen O, et al. Against the odds: what to expect in rehabilitation of chronic spinal cord injury with a neurologically controlled Hybrid Assistive Limb exoskeleton. A subgroup analysis of 55 patients according to age and lesion level. Neurosurg Focus. 2017 May;42(5):E15.

8. Khan AS, Livingstone DC, Hurd CL, et al. Retraining walking over ground in a powered exoskeleton after spinal cord injury: a prospective cohort study to examine functional gains and neuroplasticity. Journal of neuroengineering and rehabilitation. 2019 Nov 21;16(1):145.

9. Wu C-H, Mao H-F, Hu J-S, et al. The effects of gait training using powered lower limb exoskeleton robot on individuals with complete spinal cord injury. Journal of NeuroEngineering and Rehabilitation. 2018 2018/03/05;15(1):14.

10. Stampacchia G, Rustici A, Bigazzi S, et al. Walking with a powered robotic exoskeleton: Subjective experience, spasticity and pain in spinal cord injured persons. NeuroRehabilitation. 2016 Jun 27;39(2):277–83.

11. Stampacchia G, Olivieri M, Rustici A, et al. Gait rehabilitation in persons with spinal cord injury using innovative technologies: an observational study. Spinal cord. 2020;58(9):988–997.

12. Gil-Agudo Á, Megía-García Á, Pons JL, et al. Exoskeleton-based training improves walking independence in incomplete spinal cord injury patients: results from a randomized controlled trial. Journal of NeuroEngineering and Rehabilitation. 2023 2023/03/24;20(1):36.

13. Davis FD. User acceptance of information technology: system characteristics, user perceptions and behavioral impacts. International Journal of Man-Machine Studies. 1993 1993/03/01/;38(3):475–487.

14. Venkatesh V, Thong JYL, Xu X. Consumer Acceptance and Use of Information Technology: Extending the Unified Theory of Acceptance and Use of Technology. MIS Quarterly. 2012;36(1):157–178.

15. Sekhon M, Cartwright M, Francis JJ. Acceptability of healthcare interventions: an overview of reviews and development of a theoretical framework. BMC Health Serv Res. 2017 Jan 26;17(1):88.

16. Sekhon M, Cartwright M, Francis JJ. Development of a theory-informed questionnaire to assess the acceptability of healthcare interventions. BMC Health Services Research. 2022 2022/03/01;22(1):279.

17. Gorgey AS, Sumrell R, Goetz LL. 44 - Exoskeletal Assisted Rehabilitation After Spinal Cord Injury. In: Webster JB, Murphy DP, editors. Atlas of Orthoses and Assistive Devices (Fifth Edition). Philadelphia: Elsevier; 2019. p. 440–447.e2.

18. Charette C, Déry J, Blanchette AK, et al. A systematic review of the determinants of implementation of a locomotor training program using a powered exoskeleton for individuals with a spinal cord injury. Clinical Rehabilitation. 2023;37(8):1119–1138.

19. Pinelli E, Zinno R, Barone G, et al. Barriers and facilitators to exoskeleton use in persons with spinal cord injury: a systematic review. Disabil Rehabil Assist Technol. 2023 Nov 27:1–9.

20. Kinnett-Hopkins D, Mummidisetty CK, Ehrlich-Jones L, et al. Users with spinal cord injury experience of robotic Locomotor exoskeletons: a qualitative study of the benefits, limitations, and recommendations. Journal of NeuroEngineering and Rehabilitation. 2020 2020/09/11;17(1):124.

21. Manns PJ, Hurd C, Yang JF. Perspectives of people with spinal cord injury learning to walk using a powered exoskeleton. Journal of NeuroEngineering and Rehabilitation. 2019 2019/07/19;16(1):94.

22. Lizarondo L SC, Carrier J, Godfrey C, Rieger K, Salmond S, Apostolo J, Kirkpatrick P, Loveday H. Chapter 8: Mixed methods systematic reviews. In: JBI, editor. JBI Manual for Evidence Synthesis.: In: Aromataris E, Munn Z (Editors); 2020.

23. Page MJ, McKenzie JE, Bossuyt PM, et al. The PRISMA 2020 statement: an updated guideline for reporting systematic reviews. BMJ. 2021;372:n71.

24. The EndNote Team. EndNote 20 [64 bit]. Philadelphia, PA: Clarivate; 2013.

25. Veritas Health Innovation. Covidence systematic review software: Melbourne, Australia; 2023. Available from: www.covidence.org

26. McHugh ML. Interrater reliability: the kappa statistic. Biochem Med (Zagreb). 2012;22(3):276–82.

27. Lumivero. Nvivo (Version 14). 2023. Available from: www.lumivero.com

28. Hong QN, Gonzalez-Reyes A, Pluye P. Improving the usefulness of a tool for appraising the quality of qualitative, quantitative and mixed methods studies, the Mixed Methods Appraisal Tool (MMAT). Journal of Evaluation in Clinical Practice. 2018;24(3):459–467.

29. Platz T, Gillner A, Borgwaldt N, et al. Device-Training for Individuals with Thoracic and Lumbar Spinal Cord Injury Using a Powered Exoskeleton for Technically Assisted Mobility: Achievements and User Satisfaction. Biomed Res Int. 2016;2016:8459018.

30. Postol N, Spratt NJ, Bivard A, et al. Physiotherapy using a free-standing robotic exoskeleton for patients with spinal cord injury: a feasibility study. J Neuroeng Rehabil. 2021 Dec 25;18(1):180.

31. Quiles V, Ferrero L, Ianez E, et al. Usability and acceptance of using a lower-limb exoskeleton controlled by a BMI in incomplete spinal cord injury patients: a case study. Annu Int Conf IEEE Eng Med Biol Soc. 2020 Jul;2020:4737–4740.

32. Sale P, Russo EF, Russo M, et al. Effects on mobility training and de-adaptations in subjects with Spinal Cord Injury due to a Wearable Robot: a preliminary report. BMC Neurol. 2016 Jan 28;16:12.

33. Sale P, Russo EF, Scarton A, et al. Training for mobility with exoskeleton robot in spinal cord injury patients: a pilot study. Eur J Phys Rehabil Med. 2018 Oct;54(5):745–751.

34. Thomassen GK, Jørgensen V, Normann B. “Back at the same level as everyone else”-user perspectives on walking with an exoskeleton, a qualitative study. Spinal Cord Ser Cases. 2019;5:103.

35. Muijzer-Witteveen H, Sibum N, van Dijsseldonk R, et al. Questionnaire results of user experiences with wearable exoskeletons and their preferences for sensory feedback. Journal of NeuroEngineering and Rehabilitation. 2018 2018/11/23;15(1):112.

36. Lemaire ED, Smith AJ, Herbert-Copley A, et al. Lower extremity robotic exoskeleton training: Case studies for complete spinal cord injury walking. NeuroRehabilitation. 2017;41(1):97–103.

37. Kwon SH, Lee BS, Lee HJ, et al. Energy Efficiency and Patient Satisfaction of Gait With Knee-Ankle-Foot Orthosis and Robot (ReWalk)-Assisted Gait in Patients With Spinal Cord Injury. Ann Rehabil Med. 2020 Apr;44(2):131–141.

38. Heinemann AW, Jayaraman A, Mummidisetty CK, et al. Experience of Robotic Exoskeleton Use at Four Spinal Cord Injury Model Systems Centers. J Neurol Phys Ther. 2018 Oct;42(4):256–267.

39. Gagnon DH, Vermette M, Duclos C, et al. Satisfaction and perceptions of long-term manual wheelchair users with a spinal cord injury upon completion of a locomotor training program with an overground robotic exoskeleton. Disabil Rehabil Assist Technol. 2019 Feb;14(2):138–145.

40. Fundarò C, Giardini A, Maestri R, et al. Motor and psychosocial impact of robot-assisted gait training in a real-world rehabilitation setting: A pilot study. PLoS One. 2018;13(2):e0191894.

41. Evans RW, Bantjes J, Shackleton CL, et al. “I was like intoxicated with this positivity”: the politics of hope amongst participants in a trial of a novel spinal cord injury rehabilitation technology in South Africa. Disability and Rehabilitation: Assistive Technology. 2022 2022/08/18;17(6):712–718.

42. Ehrlich-Jones L, Crown DS, Kinnett-Hopkins D, et al. Clinician Perceptions of Robotic Exoskeletons for Locomotor Training After Spinal Cord Injury: A Qualitative Approach. Arch Phys Med Rehabil. 2021 Feb;102(2):203–215.

43. del-Ama AJ, Gil-Agudo Á, Pons JL, et al. Hybrid gait training with an overground robot for people with incomplete spinal cord injury: a pilot study [Clinical Case Study]. Frontiers in Human Neuroscience. 2014 2014-May-13;8.

44. Corbianco S, Cavallini G, Dini M, et al. Energy cost and psychological impact of robotic-assisted gait training in people with spinal cord injury: effect of two different types of devices. Neurol Sci. 2021 Aug;42(8):3357–3366.

45. Charbonneau R, Loyola-Sanchez A, McIntosh K, et al. Exoskeleton use in acute rehabilitation post spinal cord injury: A qualitative study exploring patients’ experiences. J Spinal Cord Med. 2022 Nov;45(6):848–856.

46. Birch N, Graham J, Priestley T, et al. Results of the first interim analysis of the RAPPER II trial in patients with spinal cord injury: ambulation and functional exercise programs in the REX powered walking aid. J Neuroeng Rehabil. 2017 Jun 19;14(1):60.

47. Benson I, Hart K, Tussler D, et al. Lower-limb exoskeletons for individuals with chronic spinal cord injury: findings from a feasibility study. Clinical Rehabilitation. 2015 2016/01/01;30(1):73–84.

48. Benson I, Hart K, Tussler D, et al. Lower-limb exoskeletons for individuals with chronic spinal cord injury: findings from a feasibility study. Clin Rehabil. 2016 Jan;30(1):73–84.

49. Scherer MJ. Assessing the benefits of using assistive technologies and other supports for thinking, remembering and learning. Disabil Rehabil. 2005 Jul 8;27(13):731–9.

50. Demers L, Monette M, Lapierre Y, et al. Reliability, validity, and applicability of the Quebec User Evaluation of Satisfaction with assistive Technology (QUEST 2.0) for adults with multiple sclerosis. Disabil Rehabil. 2002 Jan 10-Feb 15;24(1-3):21–30.

51. Day H, Jutai J, Campbell KA. Development of a scale to measure the psychosocial impact of assistive devices: lessons learned and the road ahead. Disabil Rehabil. 2002 Jan 10-Feb 15;24(1-3):31–7.

52. Zeilig G, Weingarden H, Zwecker M, et al. Safety and tolerance of the ReWalk™ exoskeleton suit for ambulation by people with complete spinal cord injury: A pilot study. The Journal of Spinal Cord Medicine. 2012 2012/03/01;35(2):96–101.

53. Esquenazi A, Talaty M, Packel A, et al. The ReWalk powered exoskeleton to restore ambulatory function to individuals with thoracic-level motor-complete spinal cord injury. Am J Phys Med Rehabil. 2012 Nov;91(11):911–21.

54. Zeilig G, Weingarden H, Zwecker M, et al. Safety and tolerance of the ReWalk™ exoskeleton suit for ambulation by people with complete spinal cord injury: a pilot study. J Spinal Cord Med. 2012 Mar;35(2):96–101.

55. Sekhon M, Cartwright M, Francis JJ. Acceptability of healthcare interventions: an overview of reviews and development of a theoretical framework. BMC Health Services Research. 2017 2017/01/26;17(1):88.

56. Barrios TA, Manley K, Dixon N, et al. Feasibility and acceptability of a remote, hands-free cognitive battery for adults with traumatic spinal cord injury. Rehabilitation Psychology. 2023;68(4):407–418.

57. Koontz AM, Garfunkel CE, Crytzer TM, et al. Feasibility, acceptability, and preliminary efficacy of a handcycling high-intensity interval training program for individuals with spinal cord injury. Spinal Cord. 2021 2021/01/01;59(1):34–43.

58. Lewis GN, Rosie JA. Virtual reality games for movement rehabilitation in neurological conditions: how do we meet the needs and expectations of the users? Disability and Rehabilitation. 2012 2012/11/01;34(22):1880–1886.

59. Unger J, Singh H, Mansfield A, et al. The experiences of physical rehabilitation in individuals with spinal cord injuries: a qualitative thematic synthesis. Disabil Rehabil. 2019 Jun;41(12):1367–1383.

60. Anderson C. Presenting and evaluating qualitative research. Am J Pharm Educ. 2010 Oct 11;74(8):141.

61. Rodgers MM, Alon G, Pai VM, et al. Wearable technologies for active living and rehabilitation: Current research challenges and future opportunities. Journal of Rehabilitation and Assistive Technologies Engineering. 2019 2019/01/01;6:2055668319839607.

62. Sumner J, Lim HW, Chong LS, et al. Artificial intelligence in physical rehabilitation: A systematic review. Artificial Intelligence in Medicine. 2023 2023/12/01/;146:102693.

63. Mitchell J, Shirota C, Clanchy K. Factors that influence the adoption of rehabilitation technologies: a multi-disciplinary qualitative exploration. Journal of NeuroEngineering and Rehabilitation. 2023 2023/06/20;20(1):80.

64. Hastall MR, Dockweiler C, Mühlhaus J, editors. Achieving End User Acceptance: Building Blocks for an Evidence-Based User-Centered Framework for Health Technology Development and Assessment. Universal Access in Human–Computer Interaction. Human and Technological Environments; 2017 2017//; Cham: Springer International Publishing.

65. Damschroder LJ, Reardon CM, Widerquist MAO, et al. The updated Consolidated Framework for Implementation Research based on user feedback. Implementation Science. 2022 2022/10/29;17(1):75.

66. Bergen N, Labonté R. “Everything Is Perfect, and We Have No Problems”: Detecting and Limiting Social Desirability Bias in Qualitative Research. Qualitative Health Research. 2020;30(5):783–792.

67. Demers L, Wessels RD, Weiss-Lambrou R, et al. An international content validation of the Quebec User Evaluation of Satisfaction with assistive Technology (QUEST). Occupational Therapy International. 1999 1999/08/01;6(3):159–175.

68. Hong QN, Gonzalez-Reyes A, Pluye P. Improving the usefulness of a tool for appraising the quality of qualitative, quantitative and mixed methods studies, the Mixed Methods Appraisal Tool (MMAT). J Eval Clin Pract. 2018 Jun;24(3):459–467.

