## Supplemental Tables for "Acceptability of Rehabilitation Exoskeleton from the Perspective of Users with Spinal Cord Injury and Healthcare Professionals: a Mixed Methods Systematic Review"

**Supplemental Table 1.** Search Strategy

| Database | Search terms | n |
|---|---|---|
| Medline (via Ovid) | (Exp Patient Acceptance of Health Care/ or Exp Patient Satisfaction/ or Attitude of Health Personnel/ or Exp Refusal to Treat/) OR (Accept* or adherence or complian* or satisfaction or utilisabilit* or "ease of us*" or intention* or opportunit* or experience* or burden or attitude* or ethical or cost* or preference* or treat* or perspective* or usabilit*).ti,ab AND "Exoskeleton Device/" or "Orthotic Devices/" OR (Exoskeleton or robotic or "Orthotic Device*" or orthose*).ti,ab. AND<br>exp "Spinal Cord Injuries"/ or exp "Spinal Cord Diseases"/ OR ("Spinal cord injur*" or "Spinal Cord Disease*").ti,ab. | 513 |
| CINAHL (via Ebsco) | (MH "Patient Compliance") or (MH "Professional Compliance") or (MH "Patient-Reported Outcomes") or (MH "Patient Satisfaction+") or (MH "Attitude of Health Personnel+") or (MH "Patient Attitudes") OR TI ( Accept* or adherence or complian* or satisfaction or utilisabilit* or "ease of us*" or intention*, or opportunit* or experience* or burden* or attitude* or ethical* or cost* or preference* or treat* or perspective* or usabilit*) OR AB ( Accept* or adherence or complian* or satisfaction or utilisabilit* or "ease of us*" or intention*, or opportunit* or experience* or burden or attitude* or ethical* or cost* or preference* or treat* or perspective* or usabilit* ) AND (MH "Exoskeleton Devices") or (MH "Orthoses") OR<br>AB ( Exoskeleton or robotic or othose* or "Orthotic Device*") OR TI ( Exoskeleton or robotic or orthose* or "Orthotic Device*" ) AND<br>(MH "Spinal Cord Injuries+") or (MH "Spinal Cord Diseases+") OR<br>TI ( ( "Spinal cord injur*" or "Spinal Cord Disease*" ) ) OR AB ( ( "Spinal cord injur*" or "Spinal Cord Disease*" ) ) | 280 |
| Embase | patient attitude'/exp or 'health personnel attitude'/exp OR<br>accept*:ab,ti or adherence:ab,ti or complian*:ab,ti or satisfaction:ab,ti or utilisabilit*:ab,ti or 'ease of us*':ab,ti or intention*:ab,ti or opportunit*:ab,ti or experience*:ab,ti or burden:ab,ti or attitude*:ab,ti or ethical*:ab,ti or cost*:ab,ti or preference*:ab,ti or treat*:ab,ti or perspective*:ab,ti or usabilit*:ab,ti AND exoskeleton (rehabilitation)'/exp OR exoskeleton:ti,ab or robotic:ti,ab or "Orthotic Device*":ti,ab or orthose*:ti,ab AND spinal cord injury'/exp or 'spinal cord disease'/de OR<br>spinal cord injur*':ab,ti OR 'spinal cord disease*':ab,ti | 490 |
| Web of Science | accept* or adherence or complian* or satisfaction or utilisabilit* OR 'ease of us*' or intention* or opportunit* or experience* or burden or attitude* or ethical* or cost* or preference* or treat* or perspective* or usabilit* AND exoskeleton or robotic or "Orthotic Device*" or orthose* AND<br>spinal cord injur*' or 'spinal cord disease*' | 776 |

**Supplemental Table 2.** Mixed Methods Appraisal Tool Checklist

| First author | Year | Citation | Screening questions: Are there clear research questions? | Screening questions: Do the collected data allow to address the research questions? | criteria1 | criteria2 | criteria3 | criteria4 | criteria5 |
|---|---|---|---|---|---|---|---|---|---|
| **Qualitative studies** | | | | | | | | | |
| | | | **1.1. Is the qualitative approach appropriate to answer the research question?** | **1.2. Are the qualitative data collection methods adequate to address the research question?** | **1.3. Are the findings adequately derived from the data?** | **1.4. Is the interpretation of results sufficiently substantiated by data?** | **1.5. Is there coherence between qualitative data sources, collection, analysis and interpretation?** | **1.1. Is the qualitative approach appropriate to answer the research question?** | **1.2. Are the qualitative data collection methods adequate to address the research question?** |
| Charbonneau | 2022 | Charbonneau, R., Loyola-Sanchez, A., McIntosh, K., MacKean, G., & Ho, C. (2022). Exoskeleton use in acute rehabilitation post spinal cord injury: A qualitative study exploring patients' experiences. The journal of spinal cord medicine, 45(6), 848–856. https://doi.org/10.1080/10790268.2021.1983314 | Yes | Yes | Yes | Yes | Yes | Yes | Yes |

**Supplemental Table 2.** Mixed Methods Appraisal Tool Checklist

| First author | Year | Citation | Screening questions | | criteria1 | criteria2 | criteria3 | criteria4 | criteria5 |
|---|---|---|---|---|---|---|---|---|---|
| | | | Are there clear research questions? | Do the collected data allow to address the research questions? | | | | | |
| Ehrlich-Jones | 2021 | Ehrlich-Jones, L., Crown, D. S., Kinnett-Hopkins, D., Field-Fote, E., Furbish, C., Mummidisetty, C. K., Bond, R. A., Forrest, G., Jayaraman, A., & Heinemann, A. W. (2021). Clinician Perceptions of Robotic Exoskeletons for Locomotor Training After Spinal Cord Injury: A Qualitative Approach. Archives of physical medicine and rehabilitation, 102(2), 203–215. https://doi.org/10.1016/j.apmr.2020.08.024 | Yes | Yes | Yes | Yes | Yes | Yes | Yes |
| Evans | 2022 | Evans, R. W., Bantjes, J., Shackleton, C. L., West, S., Derman, W., Albertus, Y., & Swartz, L. (2022). "I was like intoxicated with this positivity": the politics of hope amongst participants in a trial of a novel spinal cord injury rehabilitation technology in South Africa. Disability and rehabilitation. Assistive technology, 17(6), 712–718. https://doi.org/10.1080/17483107.2020.1815086 | Yes | Yes | Yes | Yes | Yes | Yes | Yes |

**Supplemental Table 2.** Mixed Methods Appraisal Tool Checklist

| First author | Year | Citation | Screening questions: Are there clear research questions? | Screening questions: Do the collected data allow to address the research questions? | criteria1 | criteria2 | criteria3 | criteria4 | criteria5 |
|---|---|---|---|---|---|---|---|---|---|
| Heinemann | 2018 | Heinemann, A. W., Jayaraman, A., Mummidisetty, C. K., Spraggins, J., Pinto, D., Charlifue, S., Tefertiller, C., Taylor, H. B., Chang, S. H., Stampas, A., Furbish, C. L., & Field-Fote, E. C. (2018). Experience of Robotic Exoskeleton Use at Four Spinal Cord Injury Model Systems Centers. Journal of neurologic physical therapy : JNPT, 42(4), 256–267. https://doi.org/10.1097/NPT.0000000000000235 | Yes | Yes | Yes | Yes | Yes | Yes | Yes |
| Kinnett-Hopkins | 2020 | Kinnett-Hopkins, D., Mummidisetty, C. K., Ehrlich-Jones, L., Crown, D., Bond, R. A., Applebaum, M. H., Jayaraman, A., Furbish, C., Forrest, G., Field-Fote, E., & Heinemann, A. W. (2020). Users with spinal cord injury experience of robotic Locomotor exoskeletons: a qualitative study of the benefits, limitations, and recommendations. Journal of neuroengineering and rehabilitation, 17(1), 124. https://doi.org/10.1186/s12984-020-00752-9 | Yes | Yes | Yes | Yes | Yes | Yes | Yes |

**Supplemental Table 2.** Mixed Methods Appraisal Tool Checklist

| | | | Screening questions | | | | | | |
|---|---|---|---|---|---|---|---|---|---|
| **First author** | **Year** | **Citation** | Are there clear research questions? | Do the collected data allow to address the research questions? | **criteria1** | **criteria2** | **criteria3** | **criteria4** | **criteria5** |
| Manns | 2019 | Manns, P. J., Hurd, C., & Yang, J. F. (2019). Perspectives of people with spinal cord injury learning to walk using a powered exoskeleton. Journal of neuroengineering and rehabilitation, 16(1), 94. https://doi.org/10.1186/s12984-019-0565-1 | Yes | Yes | Yes | Yes | Yes | Yes | Yes |
| Thomassen | 2019 | Thomassen, G. K., Jørgensen, V., & Normann, B. (2019). "Back at the same level as everyone else"-user perspectives on walking with an exoskeleton, a qualitative study. Spinal cord series and cases, 5, 103. https://doi.org/10.1038/s41394-019-0243-3 | Yes | Yes | Yes | Yes | Yes | No | No |
| **Non-randomized studies** | | | | | | | | | |

**Supplemental Table 2.** Mixed Methods Appraisal Tool Checklist

| First author | Year | Citation | Screening questions | | criteria1 | criteria2 | criteria3 | criteria4 | criteria5 |
|---|---|---|---|---|---|---|---|---|---|
| | | | Are there clear research questions? | Do the collected data allow to address the research questions? | | | | | |
| | | | **3.1. Are the participants representative of the target population?** | **3.2. Are measurements appropriate regarding both the outcome and intervention (or exposure)?** | **3.3. Are there complete outcome data?** | **3.4. Are the confounders accounted for in the design and analysis?** | **3.5. During the study period, is the intervention administered (or exposure occurred) as intended?** | **3.1. Are the participants representative of the target population?** | **3.2. Are measurements appropriate regarding both the outcome and intervention (or exposure)?** |
| Benson | 2016 | Benson, I., Hart, K., Tussler, D., & van Middendorp, J. J. (2016). Lower-limb exoskeletons for individuals with chronic spinal cord injury: findings from a feasibility study. Clinical rehabilitation, 30(1), 73–84. https://doi.org/10.1177/0269215515575166 | Yes | Yes | No | Yes | No | No | Yes |

**Supplemental Table 2.** Mixed Methods Appraisal Tool Checklist

| First author | Year | Citation | Screening questions | | criteria1 | criteria2 | criteria3 | criteria4 | criteria5 |
|---|---|---|---|---|---|---|---|---|---|
| | | | Are there clear research questions? | Do the collected data allow to address the research questions? | | | | | |
| Birch | 2017 | Birch, N., Graham, J., Priestley, T., Heywood, C., Sakel, M., Gall, A., Nunn, A., & Signal, N. (2017). Results of the first interim analysis of the RAPPER II trial in patients with spinal cord injury: ambulation and functional exercise programs in the REX powered walking aid. Journal of neuroengineering and rehabilitation, 14(1), 60. https://doi.org/10.1186/s12984-017-0274-6 | Yes | Yes | Yes | No | Yes | No | Yes |
| Corbianco | 2021 | Corbianco, S., Cavallini, G., Dini, M., Franzoni, F., D'Avino, C., Gerini, A., & Stampacchia, G. (2021). Energy cost and psychological impact of robotic-assisted gait training in people with spinal cord injury: effect of two different types of devices. Neurological sciences : official journal of the Italian Neurological Society and of the Italian Society of Clinical Neurophysiology, 42(8), 3357–3366. https://doi.org/10.1007/s10072-020-04954-w | Yes | Yes | Yes | No | Yes | No | Yes |

**Supplemental Table 2.** Mixed Methods Appraisal Tool Checklist

| | | | Screening questions | | | | | | |
|---|---|---|---|---|---|---|---|---|---|
| **First author** | **Year** | **Citation** | Are there clear research questions? | Do the collected data allow to address the research questions? | **criteria1** | **criteria2** | **criteria3** | **criteria4** | **criteria5** |
| del-Ama | 2014 | Del-Ama, A. J., Gil-Agudo, A., Pons, J. L., & Moreno, J. C. (2014). Hybrid gait training with an overground robot for people with incomplete spinal cord injury: a pilot study. Frontiers in human neuroscience, 8, 298. https://doi.org/10.3389/fnhum.2014.00298 | Yes | Yes | No | No | No | No | Yes |
| Fundarò | 2018 | Fundarò, C., Giardini, A., Maestri, R., Traversoni, S., Bartolo, M., & Casale, R. (2018). Motor and psychosocial impact of robot-assisted gait training in a real-world rehabilitation setting: A pilot study. PloS one, 13(2), e0191894. https://doi.org/10.1371/journal.pone.0191894 | Yes | Yes | No | Yes | Yes | No | Yes |

**Supplemental Table 2.** Mixed Methods Appraisal Tool Checklist

| First author | Year | Citation | Screening questions | | criteria1 | criteria2 | criteria3 | criteria4 | criteria5 |
|---|---|---|---|---|---|---|---|---|---|
| | | | Are there clear research questions? | Do the collected data allow to address the research questions? | | | | | |
| Kwon | 2020 | Kwon, S. H., Lee, B. S., Lee, H. J., Kim, E. J., Lee, J. A., Yang, S. P., Kim, T. Y., Pak, H. R., Kim, H. K., Kim, H. Y., Jung, J. H., & Oh, S. W. (2020). Energy Efficiency and Patient Satisfaction of Gait With Knee-Ankle-Foot Orthosis and Robot (ReWalk)-Assisted Gait in Patients With Spinal Cord Injury. Annals of rehabilitation medicine, 44(2), 131–141. https://doi.org/10.5535/arm.2020.44.2.131 | Yes | Yes | Yes | No | Yes | No | Yes |
| Platz | 2016 | Platz, T., Gillner, A., Borgwaldt, N., Kroll, S., & Roschka, S. (2016). Device-Training for Individuals with Thoracic and Lumbar Spinal Cord Injury Using a Powered Exoskeleton for Technically Assisted Mobility: Achievements and User Satisfaction. BioMed research international, 2016, 8459018. https://doi.org/10.1155/2016/8459018 | Yes | Yes | Yes | Yes | Yes | No | Yes |

**Supplemental Table 2.** Mixed Methods Appraisal Tool Checklist

| First author | Year | Citation | Screening questions | | criteria1 | criteria2 | criteria3 | criteria4 | criteria5 |
|---|---|---|---|---|---|---|---|---|---|
| | | | Are there clear research questions? | Do the collected data allow to address the research questions? | | | | | |
| Postol | 2021 | Postol, N., Spratt, N. J., Bivard, A., & Marquez, J. (2021). Physiotherapy using a free-standing robotic exoskeleton for patients with spinal cord injury: a feasibility study. Journal of neuroengineering and rehabilitation, 18(1), 180. https://doi.org/10.1186/s12984-021-00967-4 | Yes | Yes | No | No | No | No | Yes |
| Sale | 2018 | Sale, P., Russo, E. F., Scarton, A., Calabrò, R. S., Masiero, S., & Filoni, S. (2018). Training for mobility with exoskeleton robot in spinal cord injury patients: a pilot study. European journal of physical and rehabilitation medicine, 54(5), 745–751. https://doi.org/10.23736/S1973-9087.18.04819-0 | Yes | Yes | No | No | Yes | No | Yes |

**Supplemental Table 2.** Mixed Methods Appraisal Tool Checklist

| First author | Year | Citation | Screening questions | | | | | | |
|---|---|---|---|---|---|---|---|---|---|
| | | | Are there clear research questions? | Do the collected data allow to address the research questions? | **criteria1** | **criteria2** | **criteria3** | **criteria4** | **criteria5** |
| Stampacchia | 2016 | Stampacchia, G., Rustici, A., Bigazzi, S., Gerini, A., Tombini, T., & Mazzoleni, S. (2016). Walking with a powered robotic exoskeleton: Subjective experience, spasticity and pain in spinal cord injured persons. NeuroRehabilitation, 39(2), 277–283. https://doi.org/10.3233/NRE-161358 | Yes | Yes | No | No | Yes | No | Yes |
| **Quantitative descriptive studies** | | | | | | | | | |
| | | | **4.1. Is the sampling strategy relevant to address the research question?** | **4.2. Is the sample representative of the target population?** | **4.3. Are the measurements appropriate?** | **4.4. Is the risk of nonresponse bias low?** | **4.5. Is the statistical analysis appropriate to answer the research question?** | **4.1. Is the sampling strategy relevant to address the research question?** | **4.2. Is the sample representative of the target population?** |

**Supplemental Table 2.** Mixed Methods Appraisal Tool Checklist

| First author | Year | Citation | Screening questions | | criteria1 | criteria2 | criteria3 | criteria4 | criteria5 |
|---|---|---|---|---|---|---|---|---|---|
| | | | Are there clear research questions? | Do the collected data allow to address the research questions? | | | | | |
| Gagnon | 2019 | Gagnon, D. H., Vermette, M., Duclos, C., Aubertin-Leheudre, M., Ahmed, S., & Kairy, D. (2019). Satisfaction and perceptions of long-term manual wheelchair users with a spinal cord injury upon completion of a locomotor training program with an overground robotic exoskeleton. Disability and rehabilitation. Assistive technology, 14(2), 138–145. https://doi.org/10.1080/17483107.2017.1413145 | Yes | Yes | Yes | Yes | No | No | Yes |
| Lemaire | 2017 | Lemaire, E. D., Smith, A. J., Herbert-Copley, A., & Sreenivasan, V. (2017). Lower extremity robotic exoskeleton training: Case studies for complete spinal cord injury walking. NeuroRehabilitation, 41(1), 97–103. https://doi.org/10.3233/NRE-171461 | Yes | Yes | No | No | Yes | Yes | No |

**Supplemental Table 2.** Mixed Methods Appraisal Tool Checklist

| First author | Year | Citation | Screening questions | | criteria1 | criteria2 | criteria3 | criteria4 | criteria5 |
|---|---|---|---|---|---|---|---|---|---|
| | | | Are there clear research questions? | Do the collected data allow to address the research questions? | | | | | |
| Muijzer-Witteveen | 2018 | Muijzer-Witteveen, H., Sibum, N., van Dijsseldonk, R., Keijsers, N., & van Asseldonk, E. (2018). Questionnaire results of user experiences with wearable exoskeletons and their preferences for sensory feedback. Journal of neuroengineering and rehabilitation, 15(1), 112. https://doi.org/10.1186/s12984-018-0445-0 | Yes | Yes | Yes | No | No | Yes | No |
| Quiles | 2020 | Quiles, V., Ferrero, L., Ianez, E., Ortiz, M., Megia, A., Comino, N., Gil-Agudo, A. M., & Azorin, J. M. (2020). Usability and acceptance of using a lower-limb exoskeleton controlled by a BMI in incomplete spinal cord injury patients: a case study. Annual International Conference of the IEEE Engineering in Medicine and Biology Society. IEEE Engineering in Medicine and Biology Society. Annual International Conference, 2020, 4737–4740. https://doi.org/10.1109/EMBC44109.2020.9175738 | Yes | Yes | Yes | No | Yes | No | Yes |

**Supplemental Table 2.** Mixed Methods Appraisal Tool Checklist

| First author | Year | Citation | Screening questions | | criteria1 | criteria2 | criteria3 | criteria4 | criteria5 |
|---|---|---|---|---|---|---|---|---|---|
| | | | Are there clear research questions? | Do the collected data allow to address the research questions? | | | | | |
| Sale | 2016 | Sale, P., Russo, E. F., Russo, M., Masiero, S., Piccione, F., Calabrò, R. S., & Filoni, S. (2016). Effects on mobility training and de-adaptations in subjects with Spinal Cord Injury due to a Wearable Robot: a preliminary report. BMC neurology, 16, 12. https://doi.org/10.1186/s12883-016-0536-0 | Yes | Yes | No | Yes | No | Yes | Yes |
